# Effect of preventive actions and health care factors in controlling the outbreaks of COVID-19 pandemic

**DOI:** 10.1101/2020.05.09.20096255

**Authors:** Faruq Abdulla, Zulkar Nain, Md. Karimuzzaman, Md. Moyazzem Hossain, Utpal Kumar Adhikari, Azizur Rahman

**Affiliations:** Department of Statistics, Faculty of Sciences, Islamic University, Kushtia-7003, Bangladesh; Department of Biotechnology and Genetic Engineering, Faculty of Biological Sciences, Islamic University, Kushtia-7003, Bangladesh; Department of Statistics, Faculty of Mathematical and Physical Sciences, Jahangirnagar University, Savar, Dhaka-1342, Bangladesh; School of Mathematics, Statistics and Physics, Newcastle University, Newcastle upon Tyne, UK; School of Medicine, Western Sydney University, Campbelltown, NSW-2560, Australia; School of Computing and Mathematics, Charles Sturt University, Wagga Wagga, NSW-2678, Australia

**Keywords:** COVID-19 pandemic, Social distance, Lockdown, Quarantine, Case fatality rate, Life expectancy

## Abstract

With the insurgence of the COVID-19 pandemic, a large number of people died in the past several months, and the situation is ongoing with increasing health, social and economic panic and vulnerability. Due to the lack of drugs and prophylaxis against COVID-19, most of the countries are now relying on maintaining social distance as preventative actions. However, this social distancing can create global socio-economic crisis and psychological disorders. Therefore, these control measures need to have an assessment to evaluate their value in containing the situation. In this study, we analyzed the outcome of COVID-19 in response to different control measures, health care facilities, and prevalent diseases. Based on our findings, the number of COVID-19 deaths found to be reduced with increased medical personnel and hospital beds. We found 0.23, 0.16, and 0.21 as the measurement of significant non-linear relationship between COVID-19 case fatality and number of physicians *(p-value* ≤ 7.1×10^−6^), nurses and midwives *(p-value* ≤ 4.6×10^−3^), and hospital beds *(p-value* ≤ 1.9×10^−2^). Importantly, we observed a significant correlation between the reduction of COVID-19 cases and the earliness of preventive initiation. As a result, enhancing health care facilities as well as imposing the control measures in a short time could be valuable to prevent the currently raging COVID-19 pandemic. The apathy of taking nation-wide immediate precaution measure has identified as one of the critical reasons to make the circumstances worst. Notably, countries including Gambia, Nicaragua, Burundi, Namibia, and Nepal have marked in a state of danger. Interestingly, no association between the comorbidities and severity of COVID-19 was found except for few diseases including cancer, which warranted further investigation at the pathobiological level. We believe that this study could be useful in developing a control strategy in COVID-19 as well as future pandemics.

## 1. Introduction

Currently, the world has been experiencing a newly emerged pandemic causing outbreaks and global catastrophes (Huang et al., 2020; Li and De Clercq, 2020; Malik et al., 2020; Rahman and Kuddus, 2020). The etiological agent of this pandemic is a positive-sense single-stranded RNA virus designated as severe acute respiratory syndrome coronavirus 2 (SARS-CoV-2), while the disease has named as coronavirus disease 2019 (COVID-19) (Gorbalenya et al., 2020; Huang et al., 2020). The virus causes a severe form of respiratory illness, including pneumonia, fever, diarrhea, breathing difficulty, lack of smell/test, and lung dysfunctions with 4.5% of global mortality rate (Adhikari et al., 2020; Chan et al., 2020; Syal, 2020). As of April 25, 2020, COVID-19 has attributed with a total of 2,832,441 confirmed cases and has claimed 197,342 lives worldwide (https://worldometers.info/coronavirus/). Apart from the casualties, another primary concern is to deal with the ongoing global socio-economic impact caused by the COVID-19 pandemic (Gangopadhyaya and Garrett, 2020; Joo et al., 2019). By considering the spread and severity of COVID-19 with the alarming levels of global loss, the world health organization (WHO) has declared the COVID-19 situation as a pandemic and announced an event called public health emergency of international concern (PHEIC) indicating a significant threat to global health security (Bedford et al., 2020; Xu et al., 2020).

The primary source of the COVID-19 infection is infected patients as its being contagious(Chan et al., 2020; Wang and Zhang, 2020). The rate of severity and recovery from the COVID-19 is involved with the age, biological sex, and other health conditions of the infected patients (Syal, 2020). For instance, the majority of the COVID-19 patients are male (54.3%), while the fatality is more frequent (15%) in older age groups (above 80 years old) than in the younger population (Syal, 2020; Zhang et al., 2020). Furthermore, asymptomatic patients may play a critical role in the transmission process (Shen et al., 2020; Rahman and Kuddus, 2020). Moreover, studies suggest that various comorbidities (i.e., diabetes mellitus, cardiovascular, and respiratory disease) might influence the COVID-19 progression (Chan et al., 2020; Heymann and Shindo, 2020; Wang et al., 2020). Scientists are actively trying to understand the SARS-CoV-2 as well as its pathogenic nature to develop an efficient antiviral drug or vaccine (Amanat and Krammer, 2020; Dai et al., 2020; Syal, 2020). To date, however, no antiviral drugs or vaccines have found to be effective against COVID-19 (Cascella et al., 2020; Li et al., 2020).

For now, therefore, our best interest lies in subduing the symptoms/complications with available medications and controlling the viral spread through different control measures at national and global scales (Cascella et al., 2020; Hassan et al., 2020). The benefits of the data science are enormous with appropriate policy evaluation across all domains including public health and socioeconomic impacts (Rahman, 2020). In response to COVID-19, many countries have already taken such initiative such as epicenter lockdown, identifying the carrier and patients, maintaining social distances, restriction on visa and traveling, limiting public gathering, extending medical facilities, and so on (Bedford et al., 2020; Raoult et al., 2020). However, the costs of these measures for a prolonged period would be very high, resulting in an unparalleled socio-economic loss in the history of humanity. According to UNDP, the estimated revenue loss for developing countries would be at least US$220 billion (https://.covid19detectprotect.org/). Besides, domestic violence and other psychological disorders may have increased due to the prolonged period of quarantine (van Gelder et al., 2020). The actual consequences of COVID-19 remain to be observed. Some scientists also suggested developing herd immunity, an epidemiological term describing a sufficient number of immune individuals, through mass infection/vaccination (Horton, 2020). In support, some pointed out that social isolation/disconnection could heighten the risk of prevailing health conditions and mental problems (Armitage and Nellums, 2020; Gerst-Emerson and Jayawardhana, 2015; Santini et al., 2020).

Conversely, some researchers opposed to their notion regarding herd immunity. In their opinion, developing herd immunity without vaccination will result in a catastrophic outcome since the infections cover ~70% of all populations with a 0.25 - 3% case fatality rate (Kwok et al., 2020). Acquiring herd immunity will not be effective against SARS-CoV-2 due to the continuous evolution of its genome (Tang et al., 2020). Therefore, it is a high time to evaluate the effectiveness of the preventive measure in containing the situation and to value if they evaluate outweigh the costs involved. To the best of our knowledge, however, no such studies have done this yet.

In this study, we used different statistical methods to understand the relationships between the control measures and the country-wise infection index. The association of different comorbidities with the severity of the COVID-19 disease also evaluated in this limited scope. We aim to find the crucial factors in containing the situation more rationally, and suggest significant evidence-based interventions to the law enforcement personnel and policymakers accordingly. Also this study will shed light on the essential aspects that if specific comorbidities could have worsened the severity of COVID-19 and vice-versa.

## 2. Methodology

To explore the association of preventive approaches and the COVID-19 outcome, we used several secondary databases for information regarding country-wise control measures, i.e., epicenter lockdown, restriction in traveling and public services, health care measures, and many more. In addition, life expectancy, prevalence, and death rates in prevalent diseases and risk factors were considered for each country. Herein, country-based life expectancy and the COVID-19 outcome *(i.e*., confirmed cases, deaths, recoveries, and test) data has curated from the Worldometer (https://worldometers.info/). For health care measures undertaken in different countries, we employed the Humanitarian Data Exchange (https://data.humdata.org/). These measures have analyzed to check their effectiveness on the outbreak of COVID-19 by a comparative study between countries with high and low case fatality rate. Analyzing procedure involves the days calculated from the first case confirmation of a distinct nation. Also, the number of physicians, nurses, and hospital beds was obtained from the Our World in Data website (https://ourworldindata.org/).

Furthermore, we evaluated the association of life expectancy, number of physicians, nurses, and hospital beds with the COVID-19 case fatality rate. Later on, the non-linear relationships between COVID-19 case fatality rate and health care resources were measured with their respective adjusted *P*-value using nlcor R-package (Ranjan, 2020). Then, we assessed the correlation of fatal diseases with the progression of COVID-19 complications by using prevalence and death rates in several prevalent diseases and risk factors. The data regarding country-wise disease death rates and life expectancy were collected from World Life Expectancy (https://worldlifeexpectancy.com/) while prevalence rates and smoking-related statistics obtained from Our World in Data (https://ourworldindata.org/). All the data used in this study have collected on April 20, 2020.

A range of statistical and data science techniques has been used to produce results. In particular, a comparative analysis among the selected countries was done using the R graphics-package. While, a contemporary heuristic approach was utilized for estimating the various nonlinear associations between significant variables considered in this study with the R nlcor-package. All analyses for this research were completed in R Software.

## 3. Results and Discussion

### 3.1 COVID-19 and National Preventive Action

One of the principal focus of this research is to observe the effectiveness of the different preventive action plans taken by the government to control the outbreak of COVID-19. A total of 40 countries (i.e. top 20 high and top 20 low case-death countries) data have analyzed to discover the intrinsic grounds of their current conditions, as illustrated in Figure 1. In most countries among the 20 profoundly affected countries, the visa restriction policy has employed after a couple of weeks of the first COVID-19 case report, except for Iran and Switzerland. Congruently, many of the countries showed their apathy to prioritize the screening in airport and border. In contrast, the starting of declaring a state of emergency has seen distinctly by the top burdening.

**Figure 1:**
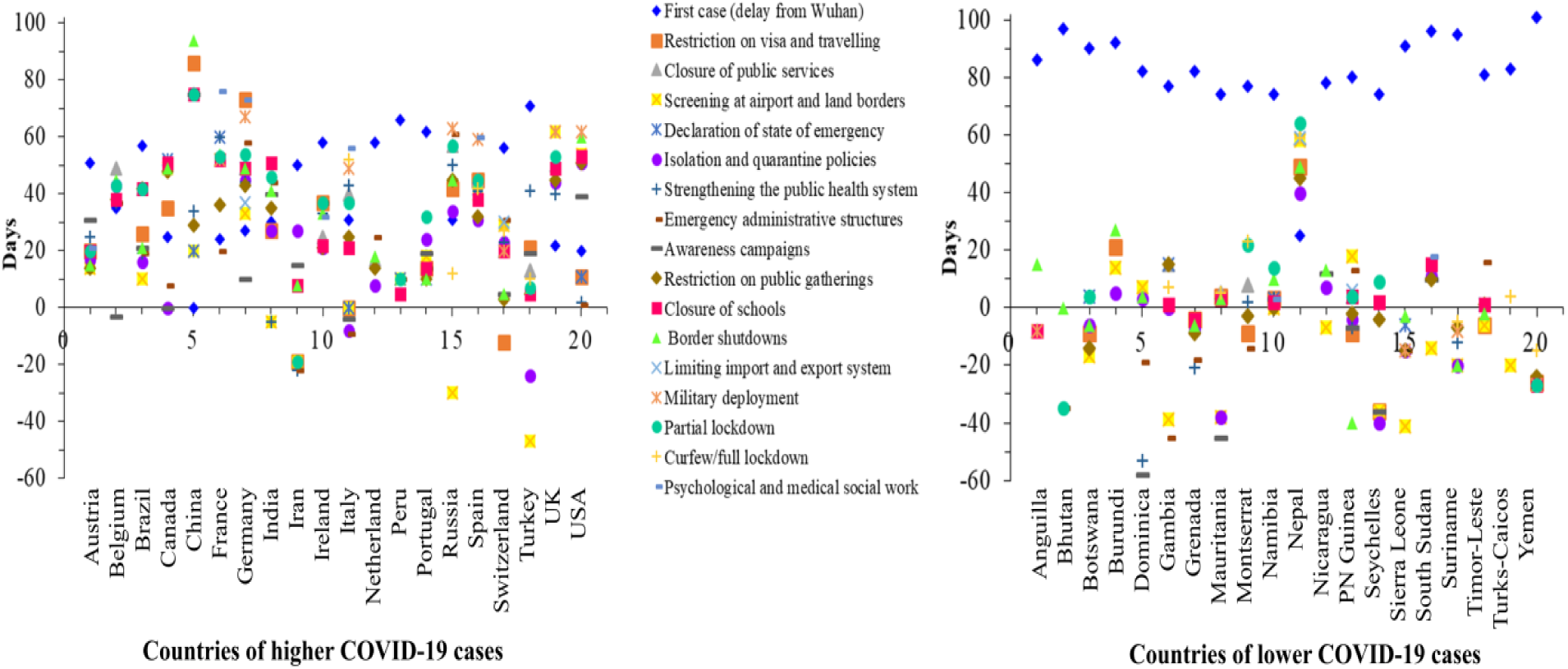
Comparative analysis of the health care measures undertaken by the government for top 20 high (left) and top 20 low (right) case-death countries.

Figure 1 also reveals that excluding the USA, Iran and India, all the top burdening nations show aloofness in the solidification of the public health structure. Among the top burdening nations USA, Italy, Iran, and Canada seem to take immediate action to stimulate emergency administrative structure compatibly, when the first confirmed cases of the disease identified. It is a scandalous finding that most of the top saddling countries introduce isolation and quarantine policies a minimum sixteen days later except few countries. Compatibly, limiting the public gathering through closing the schools, border, and public service is a splendid feature of handling the epidemic, and our findings are consistent with the findings of other research (Anderson, 2020; Cao et al., 2020; CDC, 2020a; Unicef et al., 2020). Though few countries like Iran, Turkey, Switzerland, and Peru take abrupt action to limit public gathering, deferring in other health provision care, which make their situation worse than others.

Additionally, reliable decisions such as military deployment to support the law enforcement agencies, partial lockdown, and curfew to limit public gathering have implemented latterly by most of the country, excluding Russia, Iran, and Turkey. Our findings also suggest that the scientific education-based public awareness, intense health care systems preparation, limitation of large-scale public gathering, isolation, social distancing, testing, lockdown, and other preventive measures to prevent the pandemic were effective in many societies. These findings supported by several studies those were applicable for the previous epidemic (Bedford et al., 2020; CDC, 2020b, 2020a; Kandel et al., 2007; Lau et al., 2020; Platform, 2020; Qualls et al., 2017; Xiao and Torok, 2020).

However, there is a scope of further study for identifying the real fact of spreading the disease in Iran as it takes almost every health care measure prior and immediately. On the contrary, the country with the least deaths and infections find their first infected person at minimum seventy-four days later than its origin at Wuhan, excluding Nepal. Almost all the countries found a colossal time to take immediate action and preventive measures for individuals. Most of them starting visa restriction, administrative emergency, limit public gathering, isolation policy, and partial lockdown before the identification of the first case.

Nevertheless, only a few countries make full lockdown as well as implement a testing policy in an immediate stretch. Conversely, Nepal takes almost every preventive measure lately, but still, there is a shortage of COVID-19 patients, which may indicate the successful implication of testing policy as they have done the highest number of tests. Taking account of steps implemented by different countries, this study marks a few countries as in dangerous situation based on the analysis of the preventive measure of the top twenty burdening. Among the least affected countries Gambia, Nicaragua, Burundi, Namibia, and Nepal are in a state of danger as they have taken most of the preventive measures lately than others. Only a few health care preventive measures has been taken before having the first confirmed cases by the selected countries, where they could take precautionary measures more conveniently than others as they get enough time. These marked countries do not implement the testing policy correctly though, and three of them even do not implement the full lockdown policy also, which indicates their chance to become a saddling country. Furthermore, an interesting finding of our research is that almost every country starts its awareness campaign more swiftly as the first patient identified. Still, they are in the peak of affected countries, which makes the robust validation of claiming that the scientific evidence of infection is positive to less public awareness, and it is consistent with findings from a recent study (Xiao and Torok, 2020).

### 3.2 Health Care Resources in COVID-19

The correlation between COVID-19 case fatality and health service parameters have evaluated to assess their effect on current pandemic as shown in Figure 2 (a-c). Herein, we considered three vital health system resources: (a) the number of physicians (Figure 2(a)), (b) the number of nurses and midwives (Figure 2(b)), and(c) the number of hospital beds (Figure 2(c)). All the resources have taken with an inclusion of the calculation of per 1000 peoples. It has speculated that the COVID-19 case fatality is strongly associated with these resources. We found 0.23, 0.16, and 0.21 as the measurement of non-linear relationship between COVID-19 case fatality and number of physicians, nurses and midwives, and hospital beds and these relationships are highly significant with p-value of 7.1×10^-6^, 4.6×10^-3^, and 1.9×10^-2^, respectively. As expected, the COVID-19 case fatality is high in those countries that have a lower number of physicians, hospital beds, and nurses and midwives. This observation is plausible since more doctors could treat more patients while fast health care could have provided with more nurses and hospital beds.

**Figure 2:**
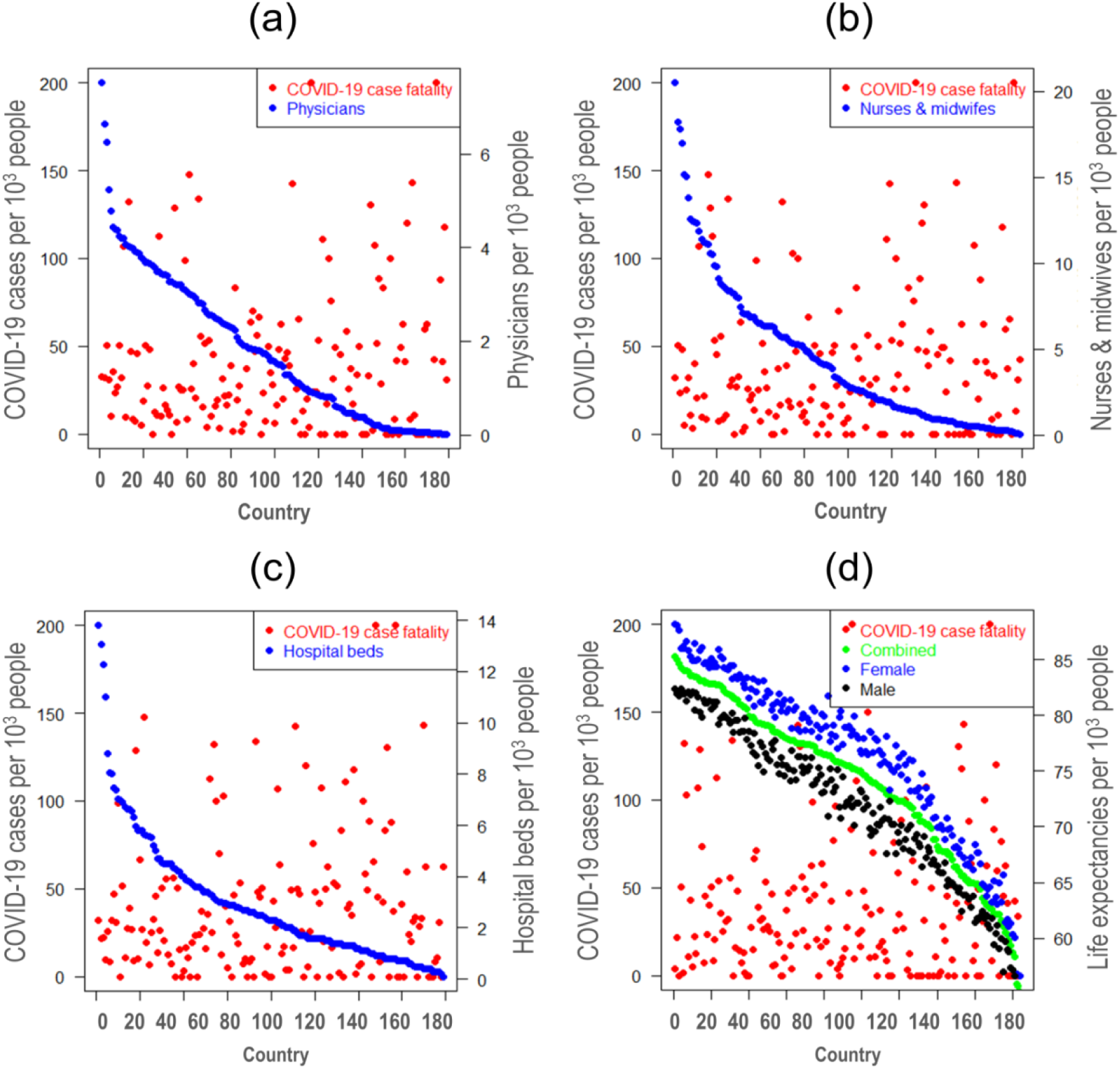
Relationship among COVID-19 case fatality, health care resources, and life expectancy where (a), (b), and (c) represent the association between COVID-19 case fatality and the number of physicians, nurses and midwives, and hospital beds, respectively while (d) indicates the relationship between COVID-19 case fatality and life expectancy by gender.

In a recent study, Ji et al. (2020) reported that the COVID-19 mortality correlates with the health care resources in China; and they concluded that the insufficient health care resources increase the COVID-19 mortality (Ji et al., 2020). In a previous study, Farahani et al. (2016) showed the association between the number of doctors, hospital beds, and nurses and midwives with the outcome of HIV-infected patients. They revealed that death from HIV would reduce by 27%, 28%, and 9% if doctors could have double, nurses can increase from 20 to 25, and five hospital beds per 10,000 people, respectively (Farahani et al., 2016).

The shortage of the selected health system resources results in the delay of care and affects the quality of care that eventually leads to more critical patients and case fatality (Drayi, 2019). For instance, Kruk et al. (2018) showed that 5 million excess deaths occurred due to the lack of quality health care in 137 low- and middle-income countries in 2016 (Kruk et al., 2018). Therefore, an increasing number of physicians, hospital beds, and nurses and midwives can significantly decrease the number of deaths from the COVID-19 pandemic.

### 3.3 Life Expectancy and COVID-19 Case Fatality

Aging is a complex cellular and molecular processes which may lead to functional and structural changes in the immune system (Sadighi Akha, 2018). Since COVID-19 is a viral disease, any changes in the immune system might influence the COVID-19 and related complications (Goronzy and Weyand, 2012; MacNee et al., 2014). Also, weak immunity has listed as one of the significant risk factors in many chronic disorders, including infections, cancer, cardiovascular, and neurodegenerative diseases (Franceschi et al., 2018; Niccoli and Partridge, 2012). Therefore, we investigated the relation between COVID-19 case fatality and the human lifespan. Due to the unavailability of the age-specific information of the COVID-19 cases and deaths, the life-expectancy was as it represents the percentage of older peoples living in a particular country.

Figure 2 (d) illustrates the relationship between life expectancy and COVID-19 case fatality rate (CFR) per 1000 cases. The CFR of COVID-19 was substantially lower in countries with higher life expectancy of both male and female. This observation is interesting as it previously thought that the CFR of COVID-19 would be higher in people older than 70 years (Glynn, 2020). The explanation of these findings could lie in their diet and lifestyle. In countries with high lifespan, people tend to have healthy food habits. For instance, a quality diet has linked to the longevity of Japanese people (Kurotani et al., 2016). The diet can positively influence the overall health and immunity that eventually lead to a long lifespan (Hale et al., 2015; Kalache et al., 2019). In a human study, for example, increasing uptake of vitamin E was found to reduce the risk of respiratory infections by 35%, especially in the upper respiratory tract (38%) where the COVID-19 initially affects (Meydani et al., 2004). Thus, our findings may provide an urgent basis to study the effect of COVID-19 on aging or vice-versa.

### 3.4 Association between COVID-19 and Comorbidities

To understand the effect of comorbidities on COVID-19, we considered country-wise death and prevalence of different diseases. Comorbidity is the prevailing conditions that can accelerate the progression and severity of certain diseases (Abood et al., 2019). For instance, several studies have reported that cardiovascular diseases, hypertension, and diabetes can increase the severity of COVID-19 (Fang et al., 2020; Wang et al., 2020). Hypothetically, there would be a positive correlation between the COVID-19 cases and these health conditions if they are associated.

Interestingly, our study reflects the opposite scenario in most cases except for a few, as shown in Figure 3.

**Figure 3:**
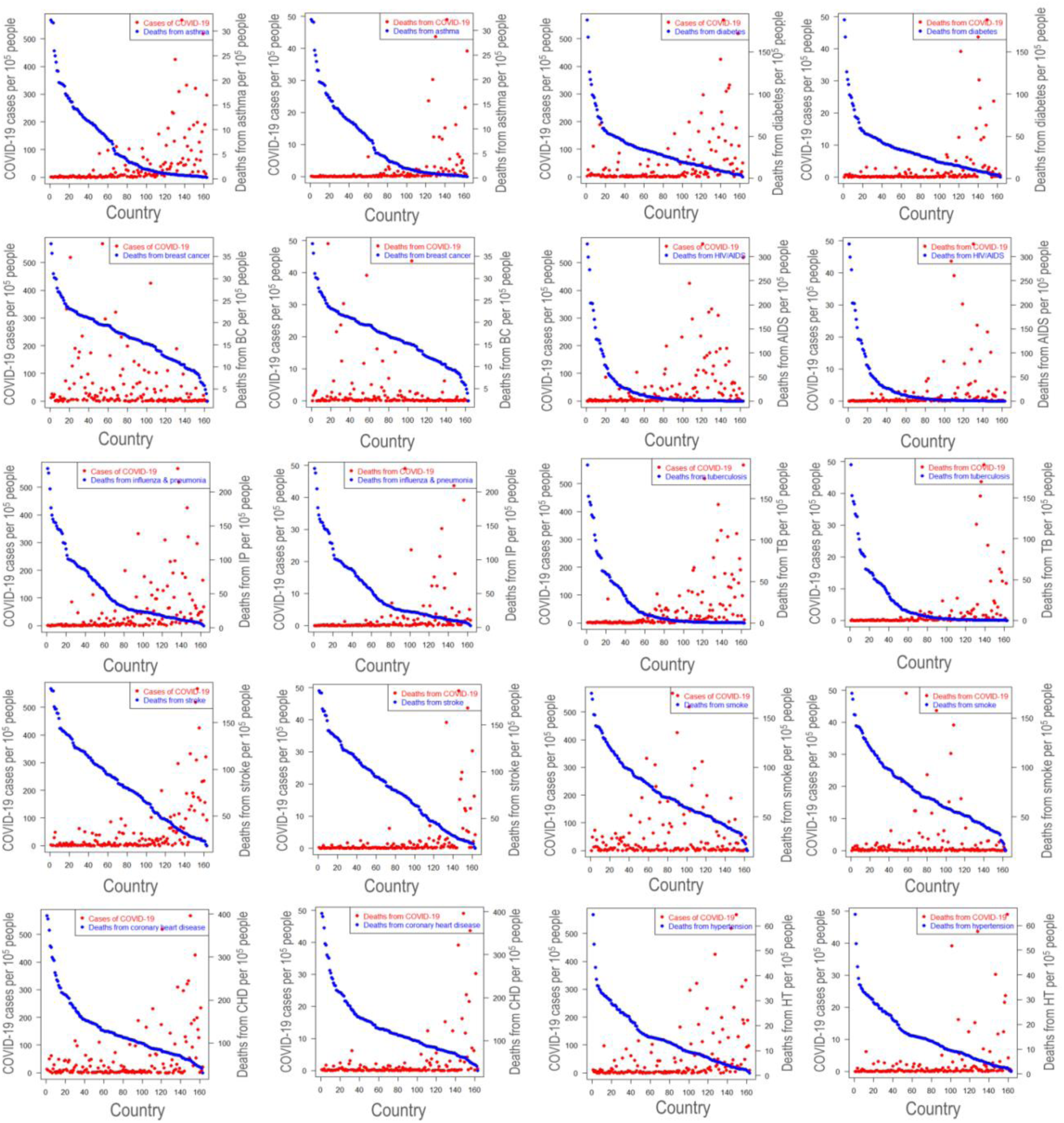
Association between COVID-19 cases and case fatality and some selected comorbidities death rates.

Notably, countries with higher death rates from asthma, cardiovascular diseases, hypertension, and diabetes showed a significantly lower amount of COVID-19 cases, which is unlike what we previously assumed. Besides, people with underlying health conditions recommended staying at home, and hopefully, they did (Kmietowicz, 2020). Moreover, people in countries where they tend to develop these disorders could be more careful and have treated with improved health services. These altogether may lead to an unusual scenario we observed in this study. However, breast cancer, leukemia, smoking, and lung cancer have found to affect the COVID-19 significantly (refer to Figure 4).

**Figure 4:**
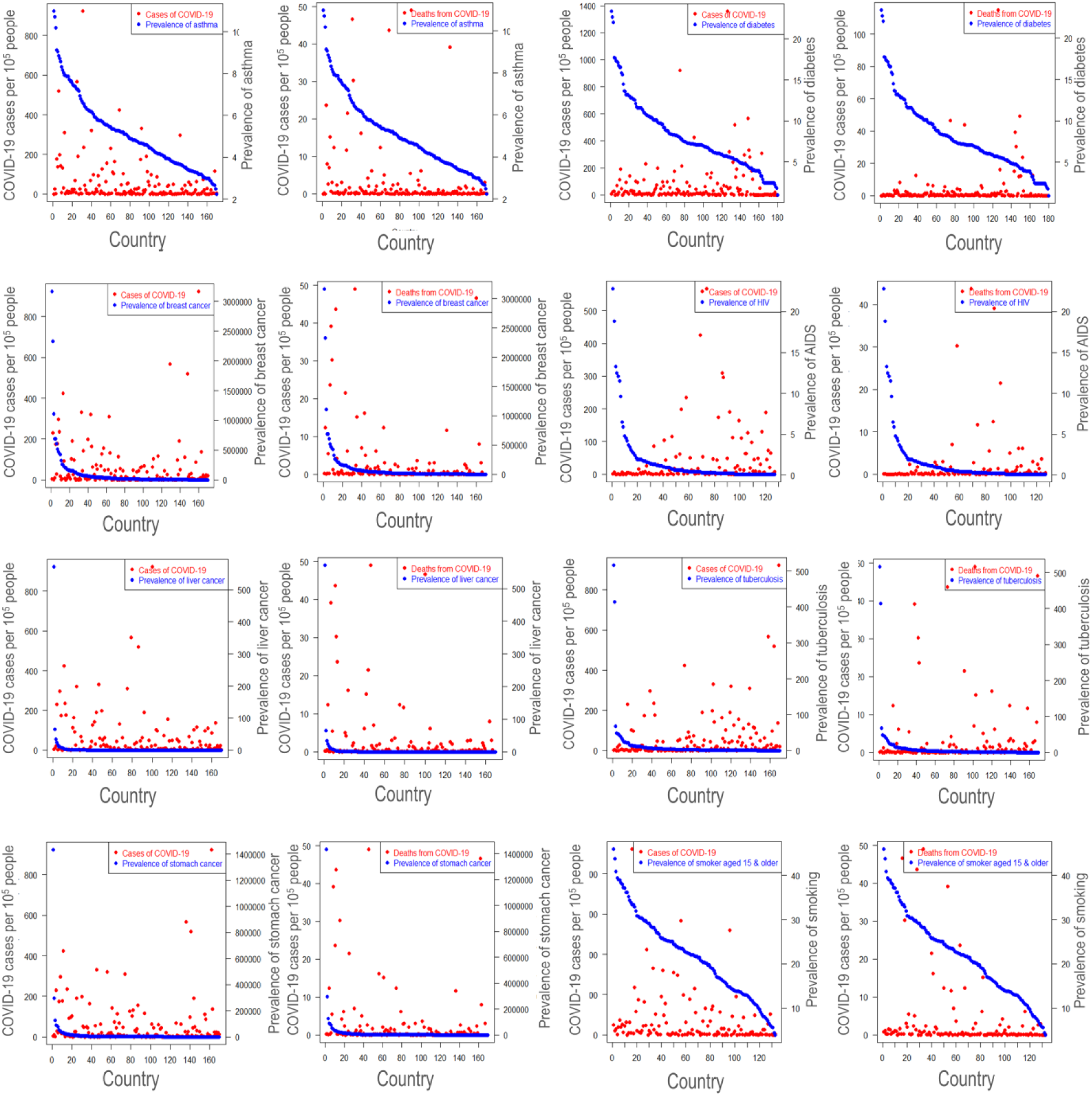
Association between COVID-19 cases and case fatality and some selected comorbidities prevalence rates.

Most importantly, the observation was quite similar when analyzed with the country-wise prevalence of several diseases. For example, countries of highly prevalent diabetes, AIDS, tuberculosis, breast cancer, and smoking showed a lower amount of SARS-CoV-2 infection (Figure 4). In the case of asthma, however, COVID-19 cases were higher in prevalent asthma countries, which are unlike the countries with higher death cases from asthma.

## 4. Conclusion

Based on the individual national risk, the immediate action of response, preparation, and readiness is an obvious step for countries that belongs to 4Cs (no cases, first cases, first clusters, and community transmission and spread). However, this study involves the scrutiny of national preventive actions, health care resources, life expectancy, and comorbidities towards the COVID-19 pandemic. Quite a few associations, correlations, as well as comparisons, have used throughout the investigations to find the relevant insights about the epidemic. The analyses of the top twenty saddling, as well as the least affected countries, have identified roughly the reasons along with the taken national health care actions. Those nations who have shown the apathy of taking immediate health care actions after the first identified case brand their circumstances more dreadful. Besides, similar lethargy of taking the immediate health care action also seen in some least burdening countries and marked as in danger. Moreover, there is an acute emergency of taking immediate health care actions to control the pandemic as most of the least saddling countries found the benefits. Also, the findings of this paper support that the COVID-19 case fatality of a country is affected by the number of physicians, hospital beds, and nurses and midwives since it has observed that there is a high COVID-19 case fatality in those countries that have a lower number of physicians, hospital beds, and nurses and midwives. The evidence of the study also validates that the increasing number of doctors, hospital beds, and nurses will result in a decline in the number of deaths from COVID-19. A stimulating finding is that the countries having higher life expectancy appear to have radically lower fatality rate in the epidemic. Correspondingly, to understand the effect of comorbidities, country-wise death and prevalence of different diseases have analyzed. Surprisingly, the results depict that the countries with higher death rates from asthma, cardiovascular diseases, hypertension, and diabetes exhibited a significantly lower amount of COVID-19 cases, which is more interesting as well as violate the pre assumptions. The authors suggest that the scientific education-based public awareness, intense health care systems preparation, limitation of large-scale public gathering, isolation, social distancing, testing, lockdown, and other preventive measures are valuable to prevent the current COVID-19 pandemic.

## Data Availability

All the data used in this study were curated from secondary data sources mentioned in the manuscript.

## Author Contributions

FA, ZN, MK, MMH and AR developed the concept and protocol; FA performed data collection and processing; FA, ZN, and MK analyzed the data; FA and ZN prepared the illustrations; FA, ZN, MK, and MMH wrote the draft manuscript with the guideline of AR; MMH, UKA and AR critically revised the manuscript; MMH and AR supervised the whole study.

## Ethical statement

Not applicable.

